# FPsim: An agent-based model of family planning

**DOI:** 10.1101/2023.02.01.23285350

**Authors:** Michelle L. O’Brien, Annie Valente, Cliff C. Kerr, Joshua L. Proctor, Navideh Noori, Elisabeth D. Root, Helen Olsen, Samuel Buxton, Guillaume Chabot-Couture, Daniel J. Klein, Marita Zimmermann

## Abstract

The biological and behavioral underpinnings of family planning (FP) unfold on an individual level, across a full reproductive life course, and within a complex system of social and structural constraints. Yet, much of the existing FP modeling landscape has focused solely on macro- or population-level dynamics of family planning. There is a need for an individual-based approach to provide a deeper understanding of how family planning is intertwined with individuals’ lives and health at the micro-level, which can contribute to more effective, person-centered design of both contraceptive technologies and programmatic interventions. This article introduces the Family Planning Simulator (FPsim), a data-driven, agent-based model of family planning, which explicitly models individual heterogeneity in biology and behavior over the life course. Agents in FPsim can experience a wide range of life-course events, such as increases in fecundability (and primary infertility), sexual debut, contraceptive choice, postpartum family planning, abortion, miscarriage, stillbirth, infant mortality, and maternal mortality. The core components of the model – fecundability and contraceptive choice, are represented individually and probabilistically, following age-specific patterns observed in demographic data and prospective cohort studies. Once calibrated to a setting leveraging multiple sources of data, FPsim can be used to build hypothetical scenarios and interrogate counterfactual research questions about the use, non-use, and/or efficacy of family planning programs and contraceptive methods. To our knowledge, FPsim is the first open-source, individual-level, woman-centered model of family planning.

**Author Summary:** Although the causes and consequences of family planning unfold on an individual level, few models of family planning consider individual heterogeneity over the life course. To that end, we introduce the methodology, parameters, and use-case(s) of the family planning simulator (FPsim). FPsim is a data-driven agent-based model of family planning, which explicitly models individual heterogeneity in biology and behaviors over a woman’s full life course to better understand the micro-level dynamics leading to more or less successful family planning programs and policies. FPsim is a data-driven model that leverages multiple sources of data to simulate realistic populations in settings that reflect real-life contexts. It is designed to be flexible and user-friendly, allowing for custom calibrations and providing integrated functions for straightforward use. This manuscript describes the model design, including its parameters, potential data sources, and limitations. We illustrate the functionality of FPsim using hypothetical scenarios that improve upon existing injectable contraceptives and introduce new injectable contraceptives into a Senegal-like setting.

## 1. Introduction

Family planning (FP) behavior and biology unfold on an individual level, across a full reproductive life course, and within a complex system of social and structural constraints. Consequences of family planning, or lack thereof, likewise unfold individually – with greater contraceptive access and use repeatedly linked to better health for women and children (Chola et al. 2015; Cleland et al. 2012; Rana and Goli 2018, 2021; Singh, Darroch, and Ashford 2014), and more empowered women (Dhak, Saggurti, and Ram 2020; Prata et al. 2017).

Yet, much of the existing FP modeling landscape has focused on macro- or population-level dynamics of family planning, with far less attention paid to individual needs and preferences (Brunson 2020; Speizer, Bremner, and Farid 2022) or individual-level consequences (Barham et al. 2021; Brunson and Suh 2020; Finlay and Lee 2018; Okenwa, Lawoko, and Jansson 2011; Schwarz et al. 2019). Due to the individual nature of the biological and behavioral underpinnings of family planning and its consequences, deeper understanding of how family planning is intertwined with individuals’ lives and health at the *micro*-level can contribute to more effective, person-centered design of both contraceptive technologies and programmatic interventions.

To that end, this article introduces the Family Planning Simulator (FPsim), a data-driven agent-based model of family planning, which explicitly models individual heterogeneity in biology and behaviors over the life course to better understand the conditions under which we might expect contraceptive decision-making to change, and, in turn, to inform programmatic and policy decision-making to expand contraceptive choice and access. To our knowledge, FPsim is the first open-source, individual-level, woman-centered model of family planning. Despite the individual nature of family planning, few FP models center individual biology and behavior. As an agent-based model, FPsim allows researchers to better understand and interrogate the individual behavioral and biological dynamics that aggregate to macro-level fertility outcomes. Integrating individual-level dynamics into the model allows for explicitly modeling interventions and programming targeted to specific groups (i.e. adolescents, postpartum women) in a heterogeneous population.

In the following sections, we outline the need for an agent-based model in the family planning field, describe the model design, data and methods used to parameterize the model, and provide illustrative examples of using FPsim for research.

### 1.1 Agent-Based Modeling

Agent-based models (ABMs; also called individual-based models) simulate realistic or theoretical populations, allowing for adaptive behavior, in which agents interact with themselves, other agents, and their environments (Railsback and Grimm 2012). ABMs link individual-level dynamics to emergent population processes, and thus have been used in social sciences and population health to address a wide range of complex issues (Billari et al. 2007; Bonabeau 2002; Grow and Van Bavel 2017; Silverman et al. 2020). An incomplete list includes such a range of demographic topics as dynamic marriage markets (Bijak et al. 2013; Billari et al. 2007); the effects of family planning efforts on conserving panda habitat in China (An and Liu 2010); sex ratio at birth (Kashyap and Villavicencio 2017); population change after armed conflict in Nepal (Williams, O’Brien, and Yao 2017, 2021); migration and mobility (Hinsch and Bijak 2022); and fertility decline and economic growth (Karra, Canning, and Wilde 2017).

### 1.2 Family Planning Modeling

Family planning biology and behavior unfold on the micro level. Fecundability, the biological capacity to conceive, is age-specific and subject to a great deal of individual variation (Smarr et al. 2017; Steiner et al. 2011; Steiner and Jukic 2016; Wesselink et al. 2017). Within households, women and couples make risk-benefit calculations at the micro-level, aligned with their preferences, desires, and intentions (Ajzen and Klobas 2013; Bongaarts and Casterline 2018; Cottingham 1997). These intentions are dynamic as families grow (Preis et al. 2020) and as women move through the life course: experiencing various states of health, reproductive outcomes, and social stability (Bledsoe, Banja, and Hill 1998; Schwarz et al. 2019).

Most agent-based models for family planning have been built to answer specific questions, e.g. helping couples with decision-making regarding delaying fertility if they have an ideal family size in mind (Habbema et al. 2015), the impact of assistive reproductive technology on fertility outcomes (Leridon 2004), the influence of son preference on sex ratio at birth (Kashyap and Villavicencio 2016), and the macro-level impact of family planning on environmental outcomes, such as panda habitat (An & Liu 2010).

Compartmental models have more commonly been used for policy and programmatic decision-making, such as Avenir Health’s Spectrum (Stover, McKinnon, and Winfrey 2010) and Impact 2 (Weinberger et al. 2012). These models have been developed as tools to understand a predefined set of *impacts* of family planning, but not necessarily to understand the dynamics driving family planning use in and of itself. A major statistical model used in FP, the Family Planning Estimation Tool or FPET (Cahill et al. 2018) projects future modern contraceptive prevalence and unmet need using historical patterns from health surveys and service statistics at the national level, but this model relies on S-curves -- which have been critiqued (Adetunji and Feyisetan 2017) -- and cannot explore deeper individual-level connections between contraception and reproductive health. To better understand *subnational* dynamics of common FP indicators, Mercer, Lu, and Proctor (2019) built a Bayesian hierarchical model that leverages spatiotemporal smoothing to integrate multiple surveys and their designs. While each of these models presents a different tool for analyzing FP questions, none provide an individual-level model that integrates the complexities of family planning dynamics – biological and behavioral – over a woman’s full reproductive life course.

## 2. Materials & Methods

### 2.1 Model Description

FPsim is an agent-based, woman-centered, data-driven model that is designed to be flexible enough to address a wide range of questions and settings. FPsim was developed in Python using the SciPy (scipy.org) ecosystem. It uses NumPy (numpy.org), Pandas (pandas.pydata.org), and Numba (numba.pydata.org) for fast numerical computing; Matplotlib (matplotlib.org) for plotting; and Sciris (sciris.org) for data structures, parallelization, and other utilities. Source code for FPsim is available via both the Python Package Index (via `pip install fpsim`) and GitHub (via fpsim.org).

### 2.2 Initialization and Parameterization

FPsim users choose a calibrated location when running the model. FPsim is currently available for Senegal, and calibrated Kenya and Ethiopia options are in development – these pre-made calibrations can be used as examples, or users can calibrate the model to their setting of choice. The model is initialized with a historical population pyramid from the context. Men and women enter the model without children and non-pregnant. Initialization of agents without history of pregnancy or childbirth creates a fictional initial cohort that will tend to have skewed outcomes. Both men and women are initialized, but men are subject to aging and mortality alone, while women can go on to experience the range of events listed in Table A1 in the appendix.

In FPsim, agents experience events and move from one state to another based on data-derived and assumption-based probabilities. Fig. 1 maps the major states and events that FPsim agents can experience. Agents are assessed for their eligibility (i.e., only pregnant women deliver; only women who are sexually active that month are eligible to conceive), and experience new events based on assigned probabilities.

**Figure 1.**
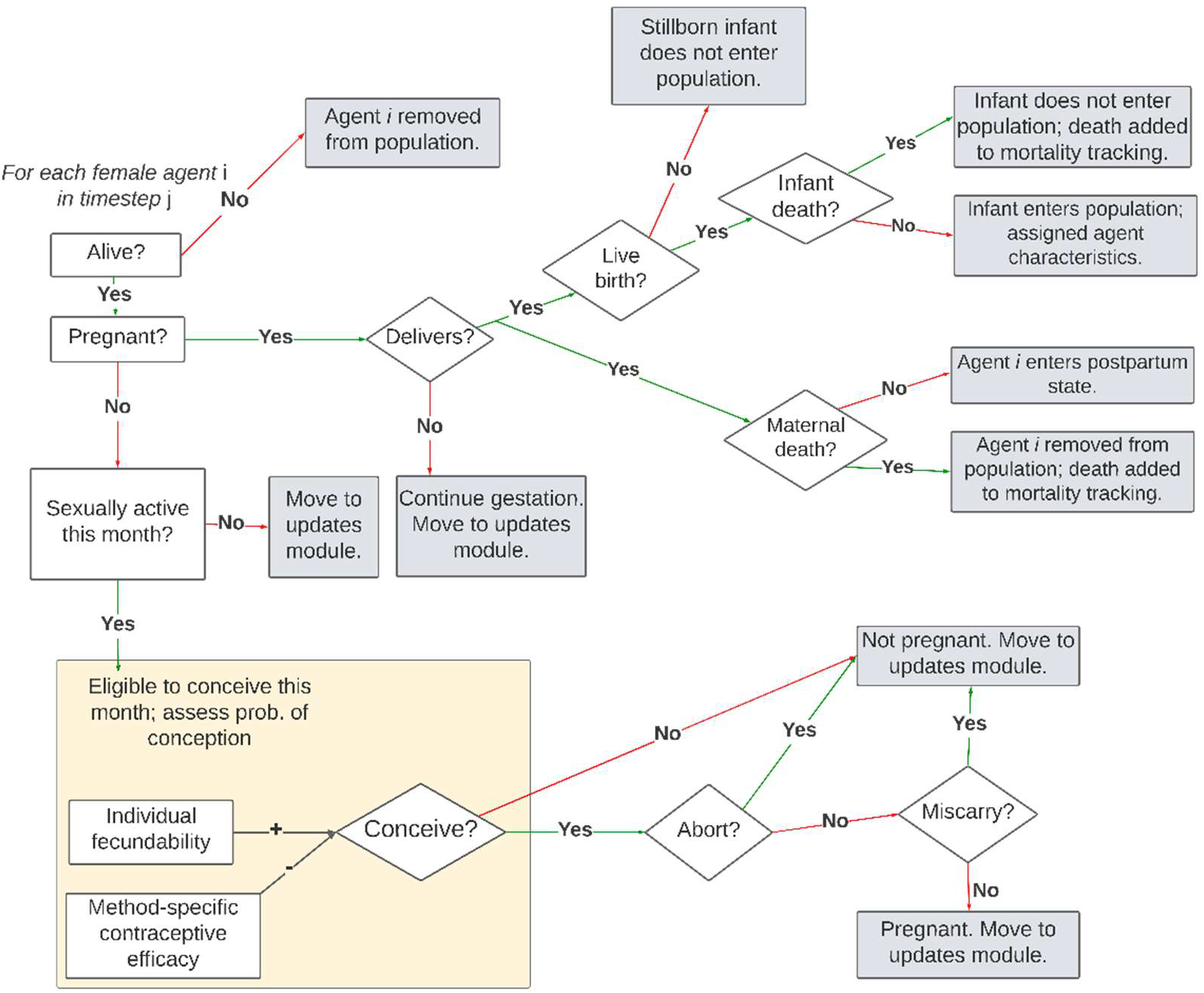
Partial map of major decisions and events encountered by FPsim agents

When the move to a new state or event is probabilistic, agents are assessed using a binomial trial – a random number between 0 and 1 is generated, and agents with an assigned probability higher than the random number will move or take that action. This allows for individual heterogeneity and, importantly for agent-based models, unpredictable behavior of some agents.

Thus, a single agent in FPsim experiences a simulated life course with probabilistic events related to her reproductive life and health. Figure 2, below, visualizes an example life course of a single FPsim-Senegal agent. How typical or atypical this agent is depends on the calibrated setting. For example, this agent may have average fertility for Senegal but higher than average for a lower fertility setting like Kenya.

**Figure 2.**
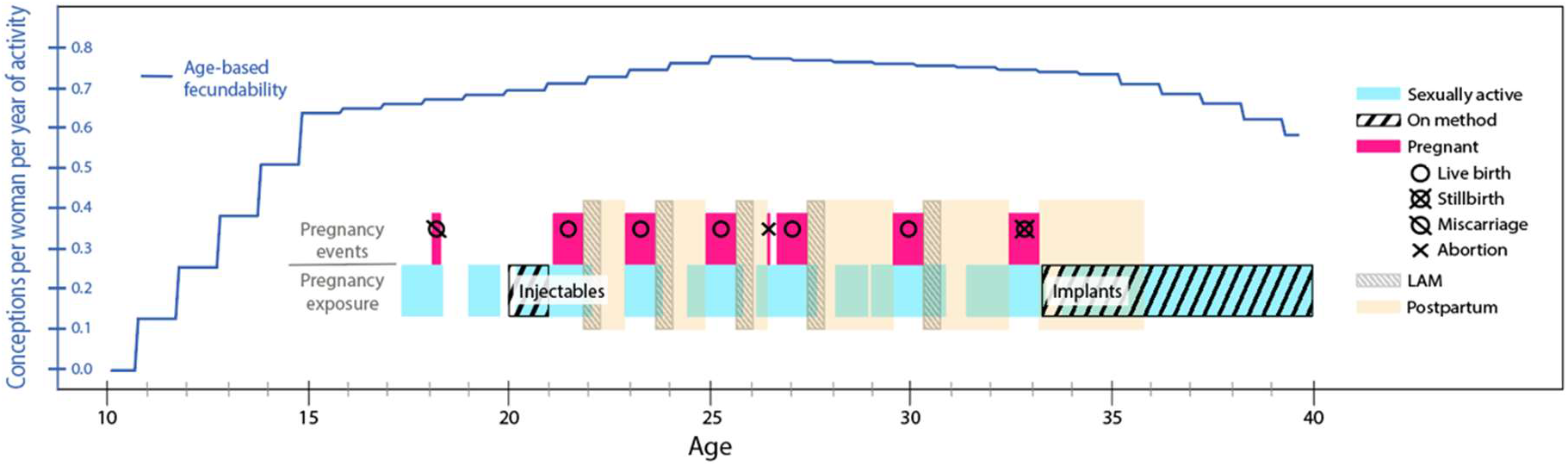
Example life course of a single FPsim-Senegal agent Note: LAM – lactational amenorrhea

**Figure 3.**
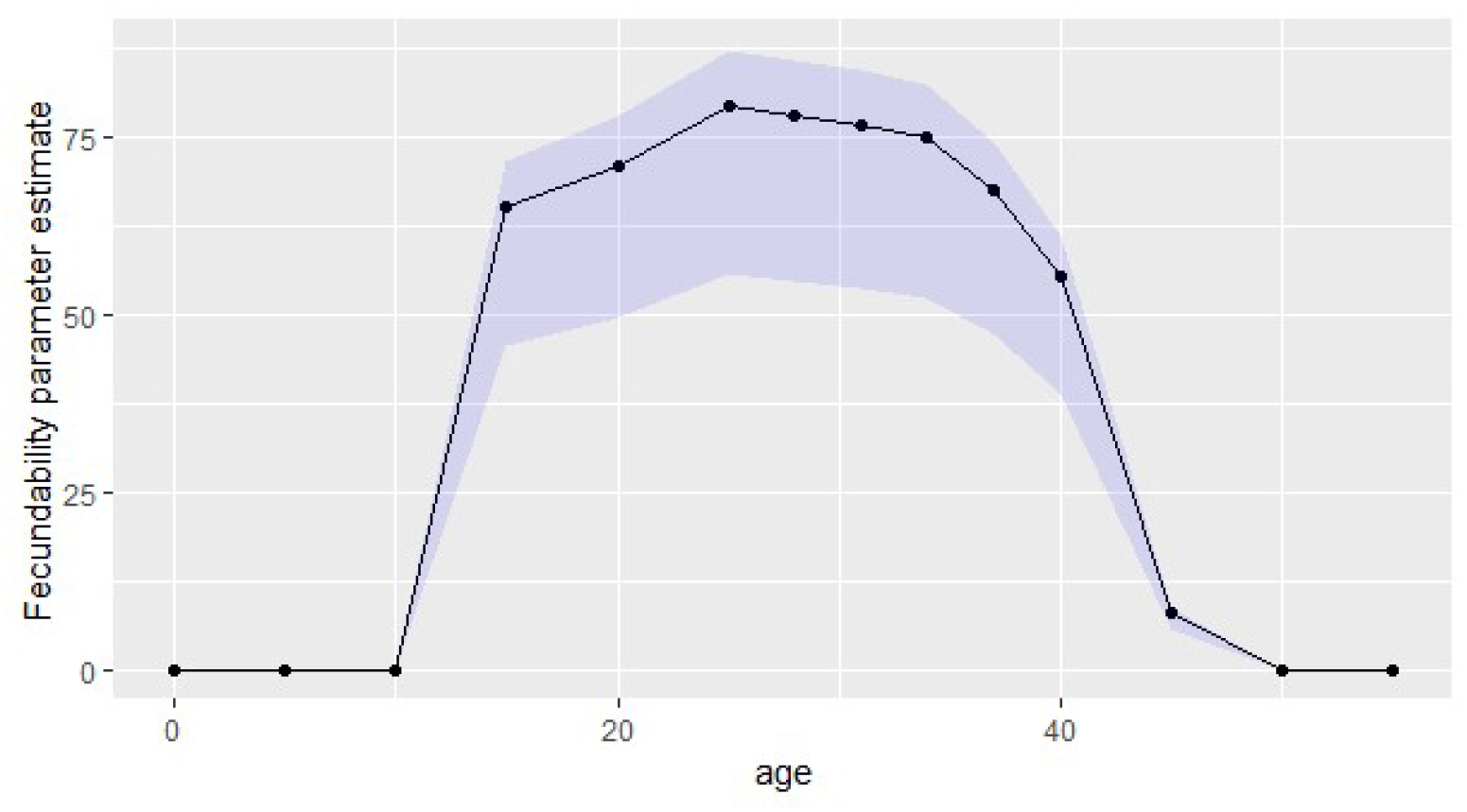
Age-specific fecundability estimates in FPsim-Senegal Shaded area represents the upper and lower bounds of individual fecundability variation.

### 2.3 Data sources & methods for parameter estimates

Depending on the parameter, agents are assigned probabilities based on a combination of any of the following: a) context-specific probabilities applied uniformly; b) age-specific probabilities; and/or c) life-stage-specific probabilities. Table A1 in the appendix lists the parameters and events that are possible for agents to experience in FPsim, as well as their respective data sources for the Senegal calibration. Note that these example data sources are meant to be informative for future calibrations, but because FPsim is data-driven and context-specific, different data sources are likely to be used for different scenarios. For instance, we parameterize matrices for initiating, discontinue, or switching contraceptive methods for Senegal using the Demographic and Health Survey (DHS) contraceptive calendars; but we use the Performance Monitoring in Action (PMA) calendars for Kenya (ICRHK 2022). Details on these matrices are in section 3.2.2. Because no one data source captures all of the intricacies and nuances within a specific country or region, FPsim calibrations are best interpreted as country- or region-like (i.e., a Senegal-like setting).

#### 2.3.1 Fecundability

Although it is commonly cited that 85% of women using no method will conceive within a year (Trussell 2011), the biological underpinnings of fertility vary over the life course, following an inverted u-shaped curve as women age. Because we simulate an entire life course, age specific fecundability estimates are critical inputs to FPsim. We parameterize fecundability as a linear interpolation of the percentage of women at each age who achieved pregnancy in the PRESTO study (Wesselink et al. 2017; Wise et al. 2015). The PRESTO study is a prospective cohort study of couples seeking pregnancies in the United States and Canada. In addition to age-specific fecundability, as women age and do not conceive, they exhibit a further decreased likelihood of conceiving (Steiner and Jukic 2016). Thus, we use additional estimates from the PRESTO study to inform a separate parameter which adjusts individual women’s fecundability downward as they age and have yet to conceive.

Women under the age of 20 are not included in the PRESTO study. Fecundability is understudied in adolescents, with rare exceptions (see, for example, Hur et al. [2020] on the relationship between undernutrition and married adolescent fecundability in Bangladesh). For the parameter in FPsim, we imputed fecundability at age 15 by applying the ratio of fertility rates for 15–19-year-olds compared to 25-year-olds. We then assume that fecundability is approximately linear from age 10 to 15 years old, as well as from age 15 to 20. The resulting distribution of age-specific fecundability estimates is shown in Fig. 2.

In addition, individuals vary widely in their fecundability. To account for this individual-level heterogeneity, we introduce a multiplier for each agent – regardless of their age-specific base fecundability or their nulliparous adjustment, we multiply their final fecundability by the individual multiplier. For the Senegal context, we use the range 0.7-1.1, as this provided the best fit for the overall population. This means that some women consistently have much lower fecundability than the PRESTO estimate for their age, and some have slightly higher.

Infertility and fecundability may vary from context to context, particularly under conditions of extreme stress (Wesselink et al. 2018). However, very few studies examine fecundability in lower- and middle-income countries, despite estimates that nearly 186 million women in LMICs experience primary or secondary infertility (Mascarenhas et al. 2012). To our knowledge, there have been no prospective cohort studies on fecundability in sub–Saharan Africa to date. Because age-specific fecundability (the biologic capacity to conceive, *not* fertility) is rarely studied in countries with DHS surveys, we use these baseline fecundability values despite their limitations.

#### 2.3.2 Contraceptive choice

Contraceptive choice in FPsim is parameterized through multiple age- and life-stage-specific choice matrices. The matrices represent the probability of switching *from* a given method, including no method, (the columns of the matrix), *to* another method, including no method (the rows of the matrix). The matrices are derived from contraceptive calendar data – in the case of Senegal, we use the Demographic and Health Survey (DHS) data from the most recent survey, and in the case of Kenya we use the longitudinal Performance Monitoring in Action (PMA) data, as the most recent DHS for Kenya is now nearly a decade old.

Agents access the annual matrices once per year, on the timestep that represents their individual birth month. Agents can only choose one method at a time, which aligns with data limitations in the field, but which does not necessarily reflect women’s concurrent usage. The choice matrices are stratified by age group, as well as postpartum status.

Nine specific methods are included: withdrawal, condoms, the pill, injectables, implants, IUDs, female sterilization, ‘other modern’ which includes emergency contraceptive and standard days method, and ‘other traditional’, which, in the DHS, encompasses any other method a respondent mentions. During their birth month, women in FPsim access the contraceptive matrices and choose a method for the year. A visual representation of one of the age-specific matrices from Senegal is shown in Figure 4.

**Figure 4.**
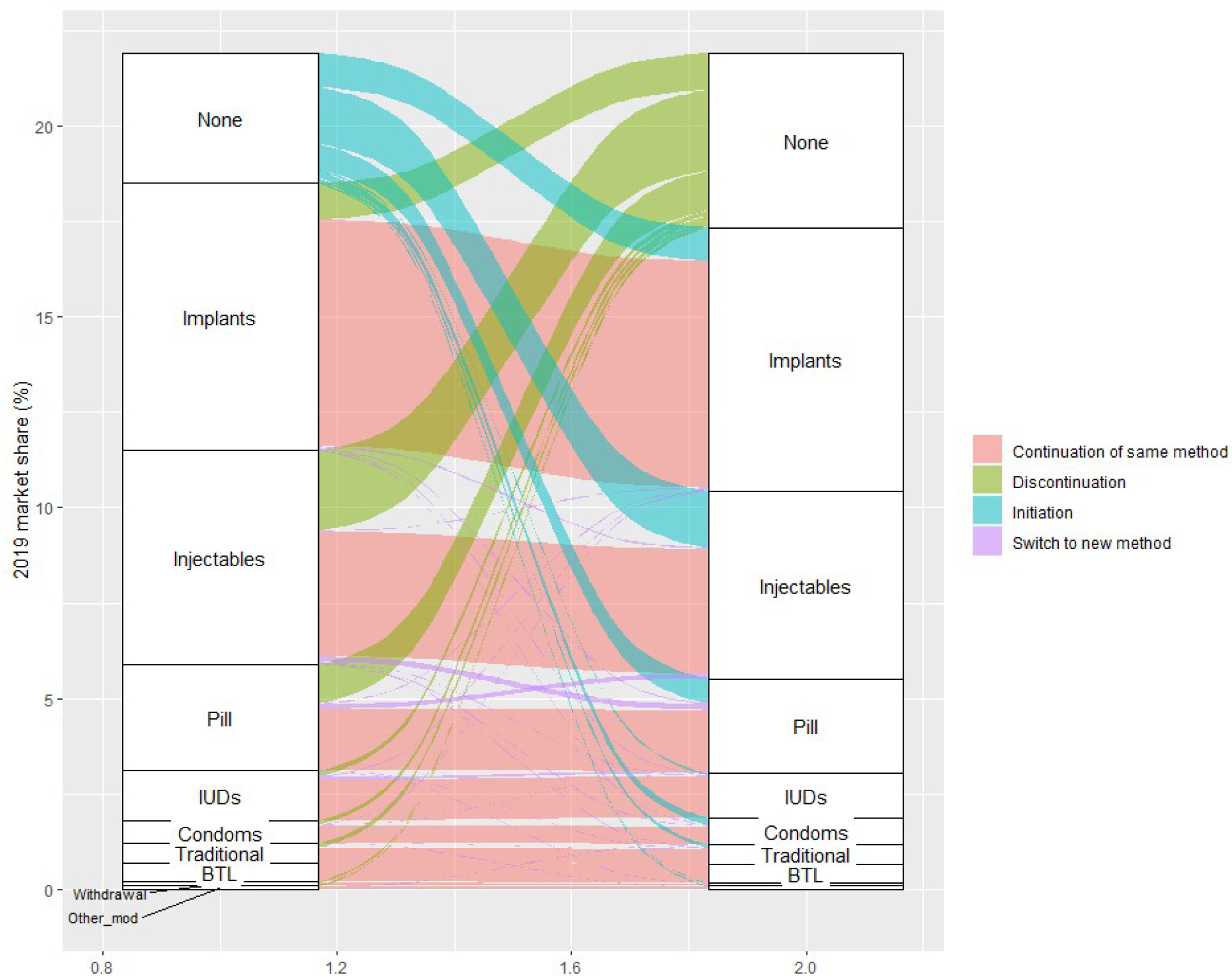
Visual representation of a switching matrix for 18–20-year-olds in FPsim-Senegal. Figure does not show ∼80% of young women in Senegal who continue non-use (None-to-None).

After delivery, separate postpartum matrices are accessed, which were derived using data from postpartum women. A one-month postpartum matrix is used to assess probability of initiating a method one month after birth, and a separate postpartum matrix is used at six months postpartum to assess the probability of starting a method or switching or discontinuing for women who initiated a method at one month postpartum. Subsequently, each woman re-enters the annual (non-postpartum) matrix at her next birth month timestep.

#### 2.3.3 Conception

One of the key life events that women experience in FPsim is conception. In any given timestep (representing one month), women who are sexually active that month are eligible to conceive. Their initial probability of conception is constituted by their individual fecundability. This probability is adjusted by their contraceptive choice – if none, there is no adjustment. All other methods are assigned an efficacy value based on failure rates in 43 DHS countries (Polis et al. 2016). Women who are within six months of delivery and exclusively breastfeeding have a probability of remaining amenorrheic and thus meeting criteria for the lactational amenorrhea method (LAM), which provides excellent protection against pregnancy (Van der Wijden and Manion 2015). We consider LAM separately from the method-specific contraceptive matrix due to these qualifying criteria.

Conception, as with all events in FPsim that are affected by multiple parameters, combines several probabilities. For instance, a 25-year-old nulliparous agent in a Senegal-like setting who was sexually active this month will have an age-specific base fecundability of 0.793 and a nulliparous (life stage) adjustment of 0.96. She has also been assigned a random variation multiplier between 0.7 and 1.1 – let us assume 0.8 for this hypothetical FPsim agent. Her total fecundability is the sum of 0.793 * 0.96 * 0.8 = 0.609. In the binomial trial to check conception, if the random number generated is lower than 0.609, this agent would conceive. However, if she also uses a modern contraceptive method, like the pill, then we apply another multiplier – (1-efficacy) – to her conception probability. In this case, her initial fecundability would be further reduced to 0.609 * (1-0.945) = 0.0335. Now this agent has greatly reduced chance (3.35%) of conception over the course of her year on the pill, which is converted to per-month probability. She will only conceive if the random number generated is lower than her individual probability in that timestep.

#### 2.3.4 Pregnancy loss and mortality

During the same timestep that agents conceive, they have a probability of terminating the pregnancy, parameterized based on Guttmacher’s context-specific abortion incidence estimates (Sedgh et al. 2015). If the pregnancy is not terminated, women may experience a miscarriage at the end of the first trimester (after three months gestation). Miscarriage probabilities are based on women’s age, where the youngest (<15) and the oldest (>35) have the highest risk. Pregnant women who are 25 years old have the lowest miscarriage probability, at 9.7 percent (Magnus et al. 2019). Once the gestation counter reaches the ninth month, women experience delivery. At the point of delivery, probabilities of stillbirth, infant mortality, and maternal mortality are assessed, in that order. Both stillbirths and infant mortality estimates follow a time-trend based on annual country-level incidence (UN Inter-agency Group for Child Mortality Estimation 2020; World Bank 2019). Adolescents under 20 years old have a higher probability of experiencing both stillbirth and infant mortality, reflected in odds ratios calculated by Noori et al. (2022).

Maternal mortality ratio (MMR) is a notoriously difficult measure to estimate with any certainty, and indeed the World Bank reports a wide 80% confidence interval in addition to their point estimates. For the Senegal calibration, we opted for published estimates of the risk of maternal death in Mali and Senegal (Huchon et al. 2013). Because those estimates are based on institutional deliveries, we provide the confidence interval to allow users to select high, medium, and low estimates of maternal mortality. The baseline estimates from Huchon et al. (2013) are then extrapolated to create a time trend based on the annual change in the World Bank indicator for MMR (WHO et al. 2019). No equivalent study of maternal death was available for Kenya, to our knowledge, and thus we use World Bank modeled estimates for Kenya’s maternal death probabilities. Because of the wide uncertainty range and the well-known issues with collecting maternal mortality data, this indicator should be interpreted with caution.

### 2.4 Data gaps and assumption-based parameters

One of the most informative aspects of a data-driven agent-based model is that researchers are forced to precisely indicate and quantify relationships between agents, agent history, and the agent’s environments. In doing so, the model development itself can highlight critical data gaps in the field. Insights into those critical data gaps can inform investments in data collection and programs. Although we leverage multiple data sources in FPsim, we do identify data gaps, for which we have used assumption-based parameters.

One of the most impactful of these assumptions, *exposure*, is a multiplier applied directly to conception probabilities, based on women’s age and/or parity, to proxy residual exposure to pregnancy not captured by data. For the Senegal calibration, we found that using age-based exposure was not necessary. However, we did find it necessary to use parity-based exposure to reduce the likelihood of conception once women reach parity 7 and above. In contexts with scarce data, researchers may find that exposure corrections, especially at the margins, are necessary to achieve realistic pregnancy outcomes.

Birth spacing patterns present a unique challenge to simulating synthetic cohorts. In part, this is due to mismatching data timelines for most demographic surveys (including the DHS). Contraceptive calendar data typically captures 1-5 years retrospectively, but shouldn’t be considered reliable much more than 12-24 months due to recall bias (Callahan and Becker 2012). On the other hand, fertility data, in which respondents typically report birth month and year of each living child, spans a woman’s entire reproductive life course up to the time of interview. This creates a gap in knowledge where researchers can identify birth spacing between births for which we do not know that woman’s contraceptive use or non-use. Because of this gap, we developed a birth spacing preference parameter, which increases or decreases an agent’s likelihood of being sexually active while she is postpartum. This parameter indirectly impacts her likelihood of conception, via her eligibility each timestep.

## 3. Results

### 3.1 Using FPsim for Research

Given a set of basic biological constraints (fecundability, conception, pregnancy), we can use FPsim to model the impact of dynamic individual-level decisions about contraceptive use and/or shifting probabilities of pregnancy-related events (e.g., abortion) on specified metrics over time. How we use FPsim depends on what kind of information we have, and generally fall under one of two categories: data-driven research questions, and assumption-based research-questions.

Data-driven research questions leverage additional data sources, including historical datasets, user insights and market research, and so on, to inform how we anticipate behavioral changes for some women. For instance, some research questions might investigate the compounding effects over time of increased uptake of a specific method. With FPsim, we could also examine those effects if the changes are limited to specific age groups, or postpartum women. We can investigate how switching behaviors impact the roll-out of a new contraceptive method. For example, a researcher may want to compare scenarios in which we roll out a new injectable and a new implant in a Senegal-like setting where contraceptive prevalence is low; but amongst method users, injectables and implants are already popular. We could examine how investing in an improved injectable or implant might impact the method mix and other family planning outcomes. Identifying and quantifying data gaps would also fall under data-driven research questions.

Assumption based research, on the other hand, implements user-defined assumptions into the model to reach particular outcomes. The researcher may ask, for example, what magnitude of behavioral change (uptake) would have needed to occur to meet FP2020 mCPR goals in a certain country? Another example of an assumption-based question would be what kind of gains in adolescent postpartum family planning would need to occur to reduce rapid repeat pregnancies, and in turn, adolescent maternal mortality.

### 3.2 Scenarios

To examine any research question using FPsim, the model needs to be calibrated to a setting, either a pre-programmed setting like Kenya or Senegal, or a custom calibration that users create for their own research questions. Once the model is calibrated, custom intervention scenarios can be built to investigate a wide range of research questions. Users can adjust nearly any parameter with the built-in scenarios script, including abortion, the probabilities of adverse outcomes (e.g. stillbirth), and the dimensions of any existing method (such as efficacy, initiation, switching to another method, and discontinuation). Users can also add a new contraceptive technology by adding in a row and method to the existing contraceptive matrices. These scenarios can be built to affect an entire population, or they can be written to affect specific sub-populations, which may be defined by characteristics including age, parity, or postpartum status.

FPsim allows for straightforward, user-friendly scenario-building. The following example scenarios are intended to illustrate the mechanics of building scenarios in FPsim, rather than to answer any one specific research question. In the first example, we build hypothetical scenarios for FPsim-Senegal, in which we 1) increase the efficacy of existing injectable methods to 99%; 2) double the probability of all women initiating injectables; 3) double the probability of women over 35 initiation injectables; and 4) combine scenarios 1 and 3, increasing the efficacy of injectables for all, and doubling the initiation for women over the age 35. Figure 6 displays the code snippet used to build the scenarios.

**Figure 6.**
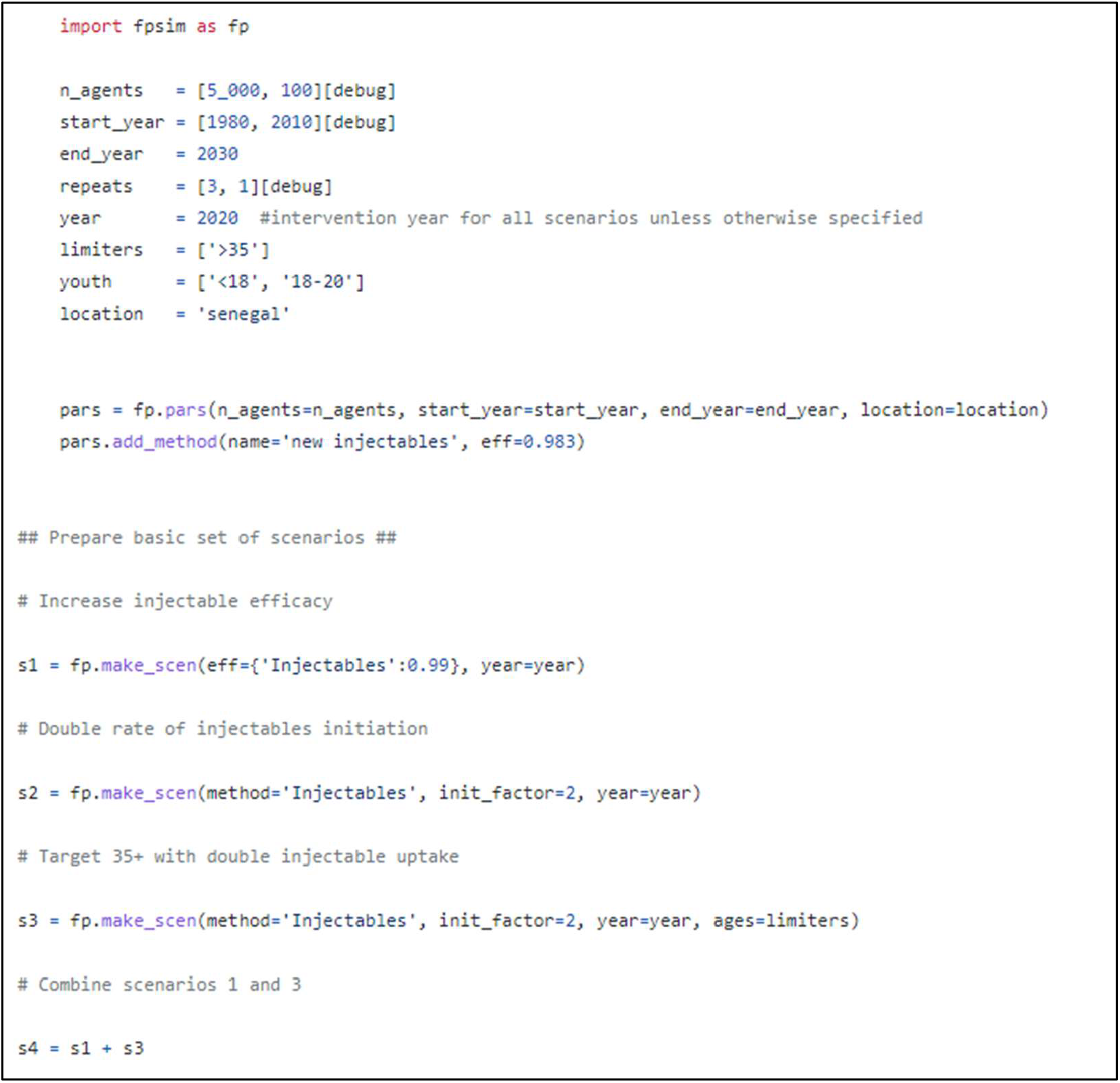
Basic scenario building in FPsim

Full python scripts to replicate the scenarios and outputs of these sample scenarios are publicly available at: https://github.com/fpsim/fpsim_technical.

More complex scenarios, to add in a new contraceptive technology, for instance, take a slightly different shape. In the example below, we are building a single scenario in three distinct parts. Figure 7, below, contains the code snippet to build this three-part scenario.

**Figure 7.**
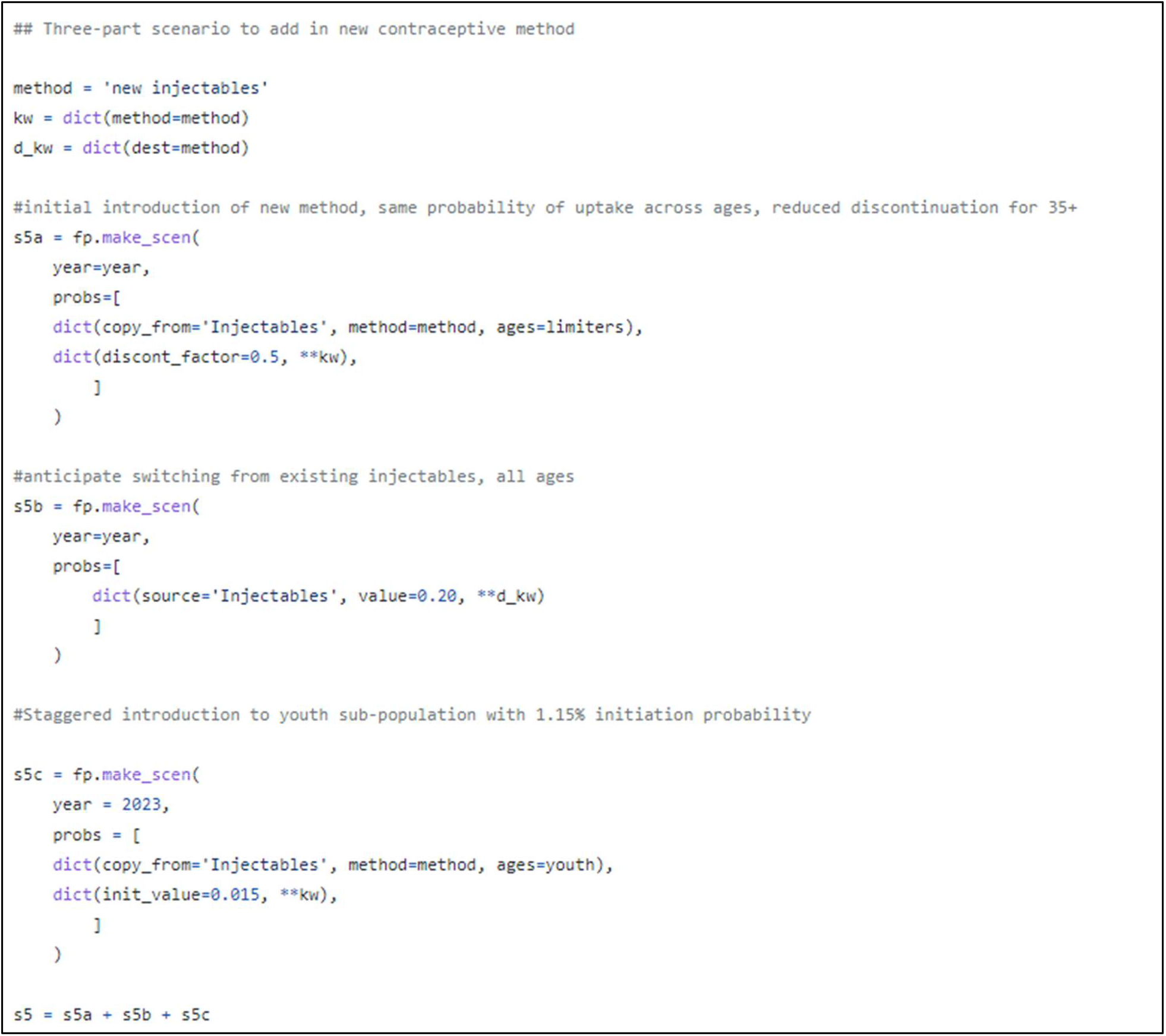
Complex scenario building with FPsim

In part a, we first introduce the new injectable method by copying over characteristics from the existing injectable (`dict(copy_from=‘Injectables’, method=method, ages=limiters)`). We then identify the characteristics we want to change for the named age group, limiters, who are over 35. In this case, we want to halve the discontinuation probability for the newly introduced injectables (`discont_factor=0.5`), perhaps assuming the new injectables will address issues like side effects and be more appealing to women than the existing method. In part b, we anticipate 20% probability of switching from existing injectables to the new injectables. In part c, we add in a staggered introduction that focuses on the youth (under 20) population. As before, we simply add up the three scenario parts to incorporate all aspects of our new contraceptive technology into a single scenario.

### 3.3 Output

FPsim has integrated plot options that can be utilized after a single sim or after a multiple simulation run with user-defined scenarios. The default plotting option (`plot()`) includes mCPR, a cumulative count of live births, stillbirths, maternal deaths, and infant deaths, and the infant mortality rate. Fig. 8, below, shows the default output for our basic set of sample scenarios.

**Figure 8.**
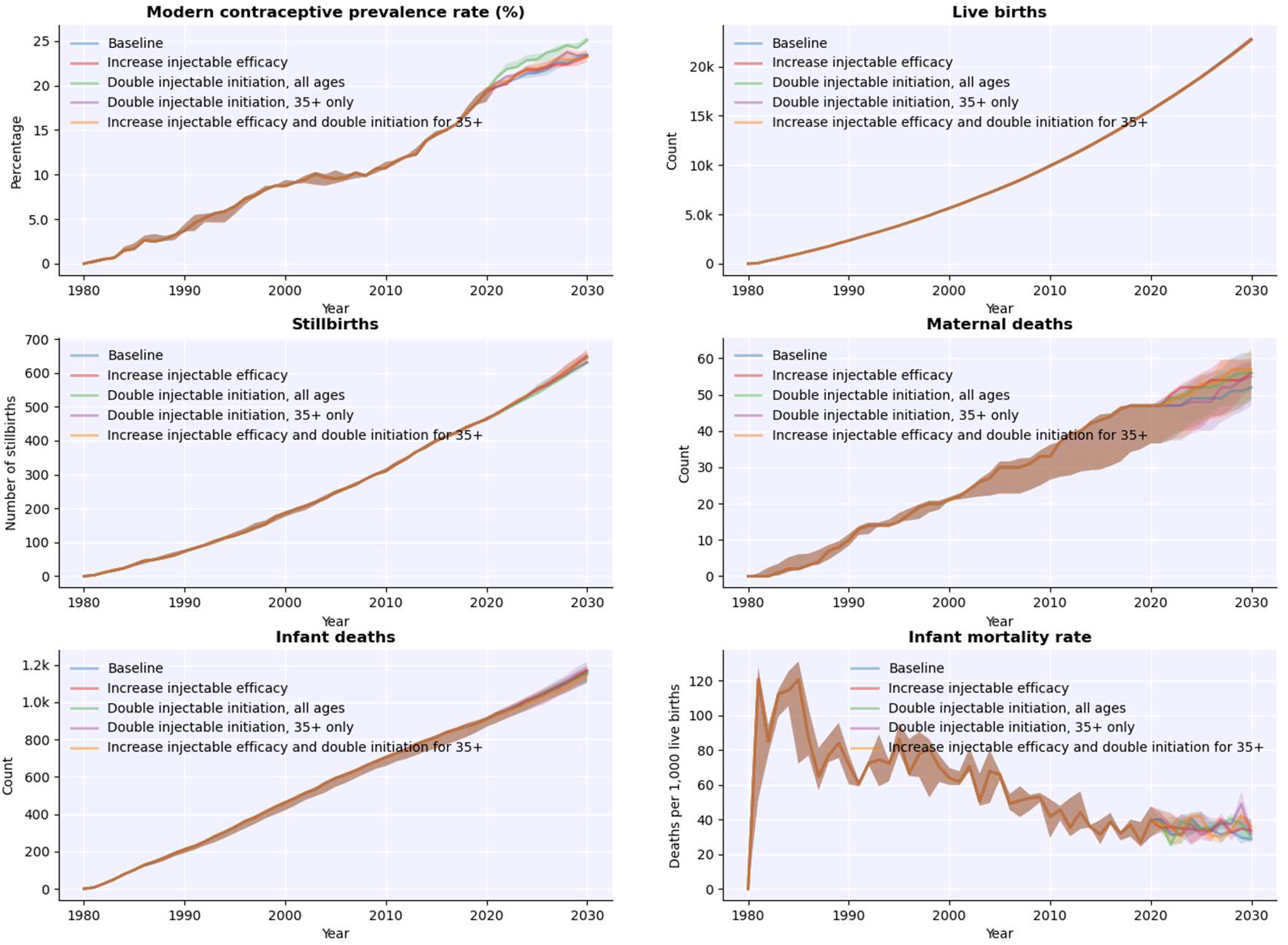
Default output after running scenarios in FPsim

Other integrated plotting options include adverse pregnancy outcomes, multiple definitions of contraceptive prevalence, and method mix over time. Fig. 9 shows the method mix plotting of our more complex scenario, in which we may want to track the how the switching patterns we identified impact the existing injectables (light blue) when we introduce the new injectables (salmon).

**Figure 9.**
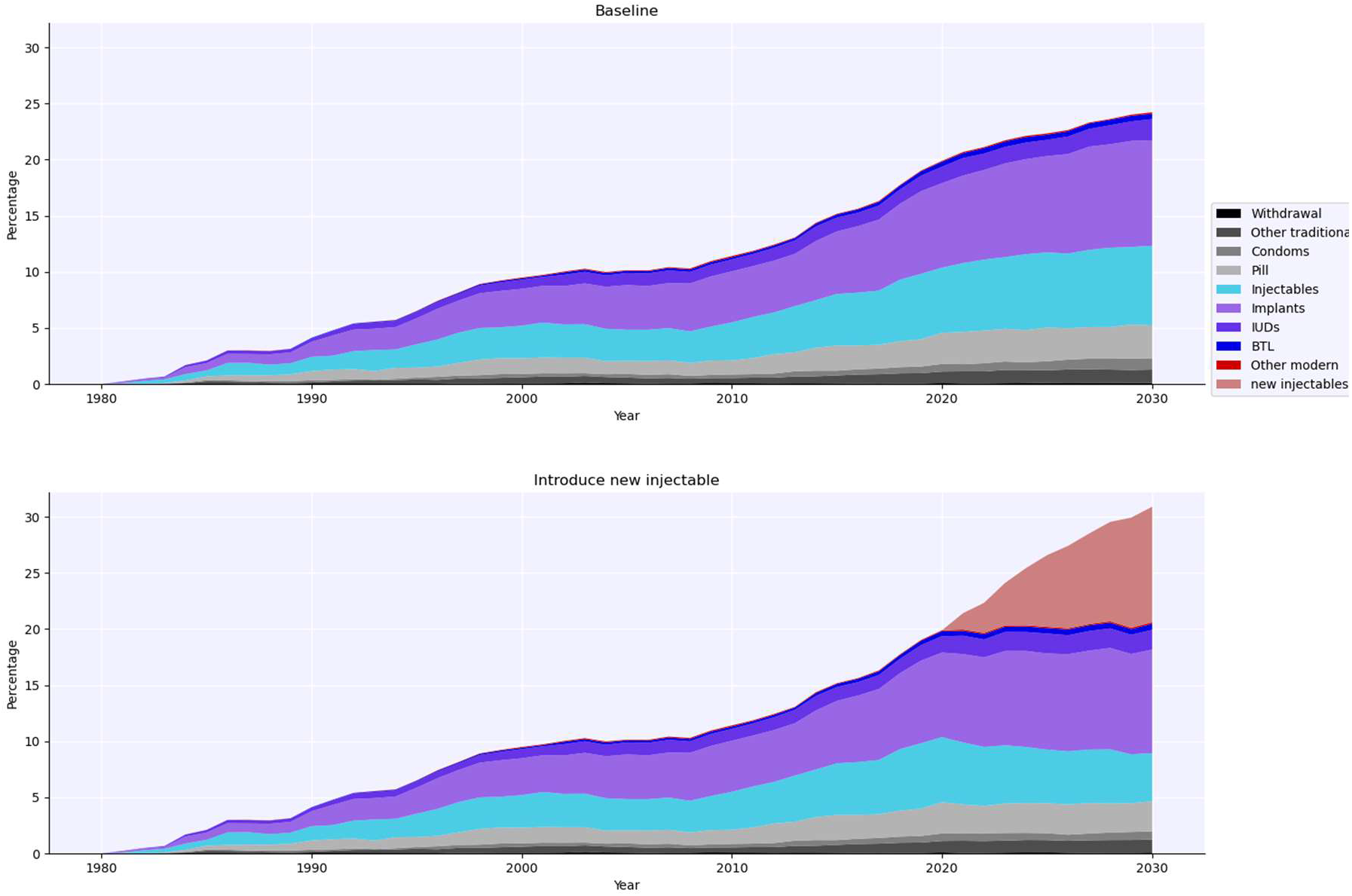
Method mix plotting to track the introduction of a new method in FPsim

## 4. Discussion

Although there currently exist FP models that provide cross-country comparisons, we see a need for an FP model designed to explicitly consider individual trajectories, to complement and augment our understanding of the conditions under which family planning programs succeed or fail. With a focus on macro-level inputs and outputs alone, we risk missing the individual heterogeneity in biology and behavior, that underlie and can deeply impact family planning dynamics at both the micro- and macro-levels. We designed Family Planning Simulator (FPsim), which centers a woman’s individual life course. This allows FPsim to generate insights into how individual behavior and biology impact fertility and health outcomes, including contraceptive prevalence, pregnancy loss and mortality, and method mix. With individual-level modeling and a life course perspective, we can better capture how probabilistic behaviors interact with biology, and how events and activities impact women differently throughout their life. FPsim provides an agent-based environment in which researchers can leverage multiple sources of data and interrogate assumptions. Researchers and policymakers alike can use the tool to improve goal setting through examining behavioral change – i.e. changing demand – rather than relying on supply-side factors alone. FPsim’s flexible and modular simulation scenarios provide an opportunity to explore the impact of policies and investments in family planning using a modeling tool designed around a woman’s unique reproductive life course.

As with any computational modeling methodology, FPsim has its limitations. No simulation model can replace rigorous data collection and analysis. The insights that FPsim produces are only as good as the inputs and expertise that inform the model. FPsim is best used as a tool to ask questions about what could be, or what could have been, using counterfactual scenarios informed by robust data and expert opinion. With these caveats, we aim for FPsim to be a user-friendly and informative tool that can supplement the family planning research and evaluation landscape to help decision-makers better target the most impactful investments, programming, and implementation strategies.

## Data Availability

All data & output produced in the manuscript are available online at https://github.com/fpsim/fpsim_technical

https://github.com/fpsim/fpsim

## Appendix

**Table A1.**
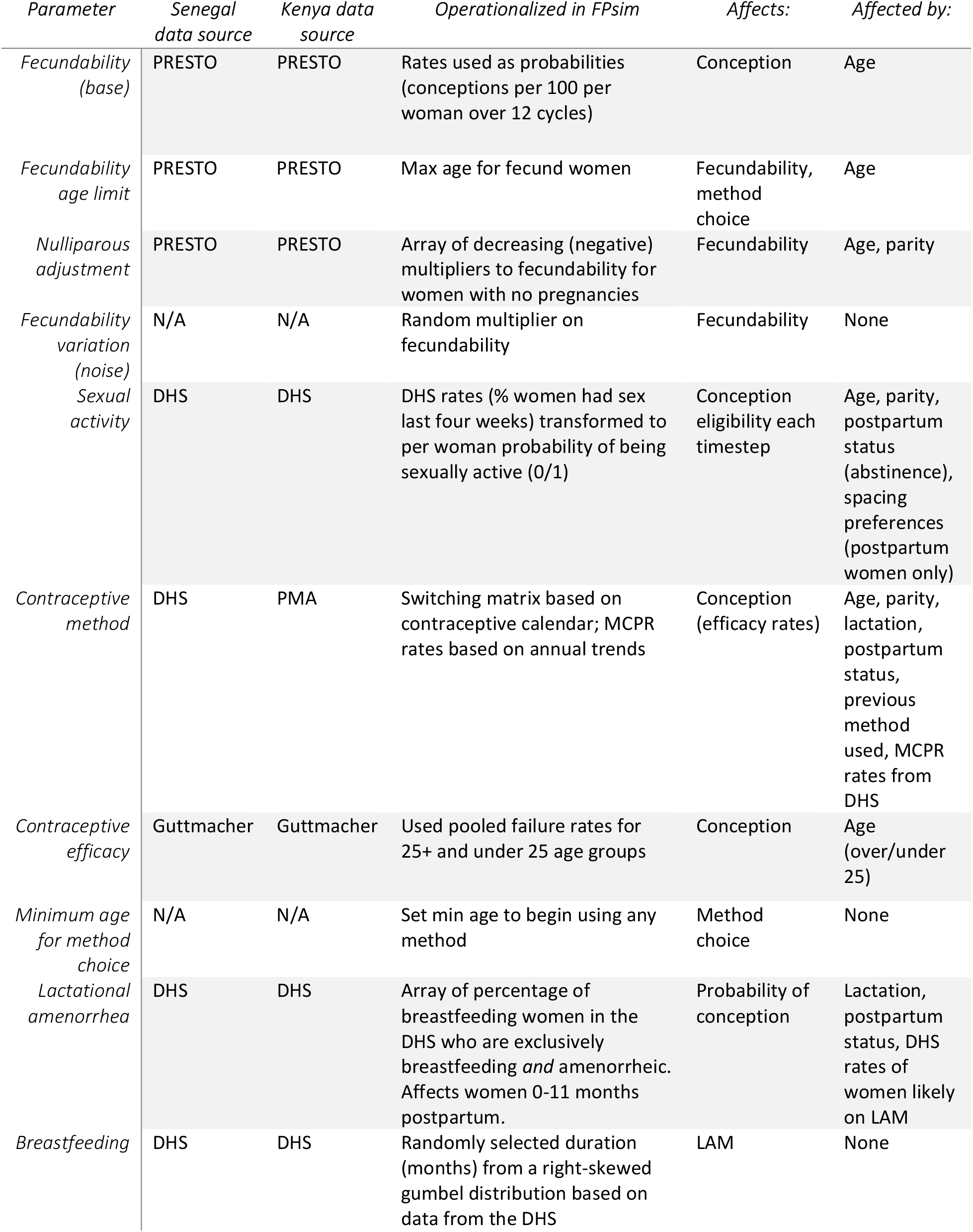
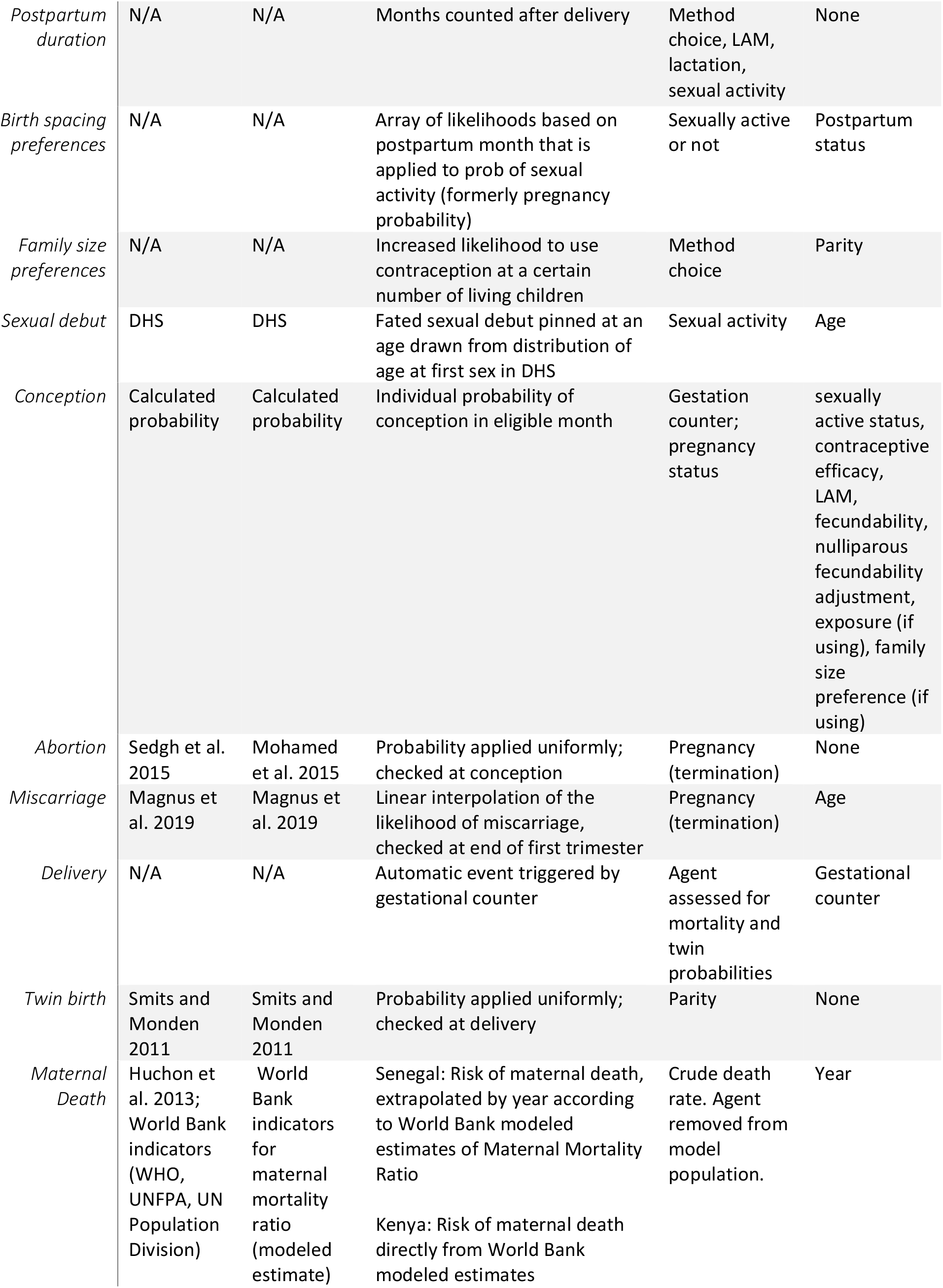
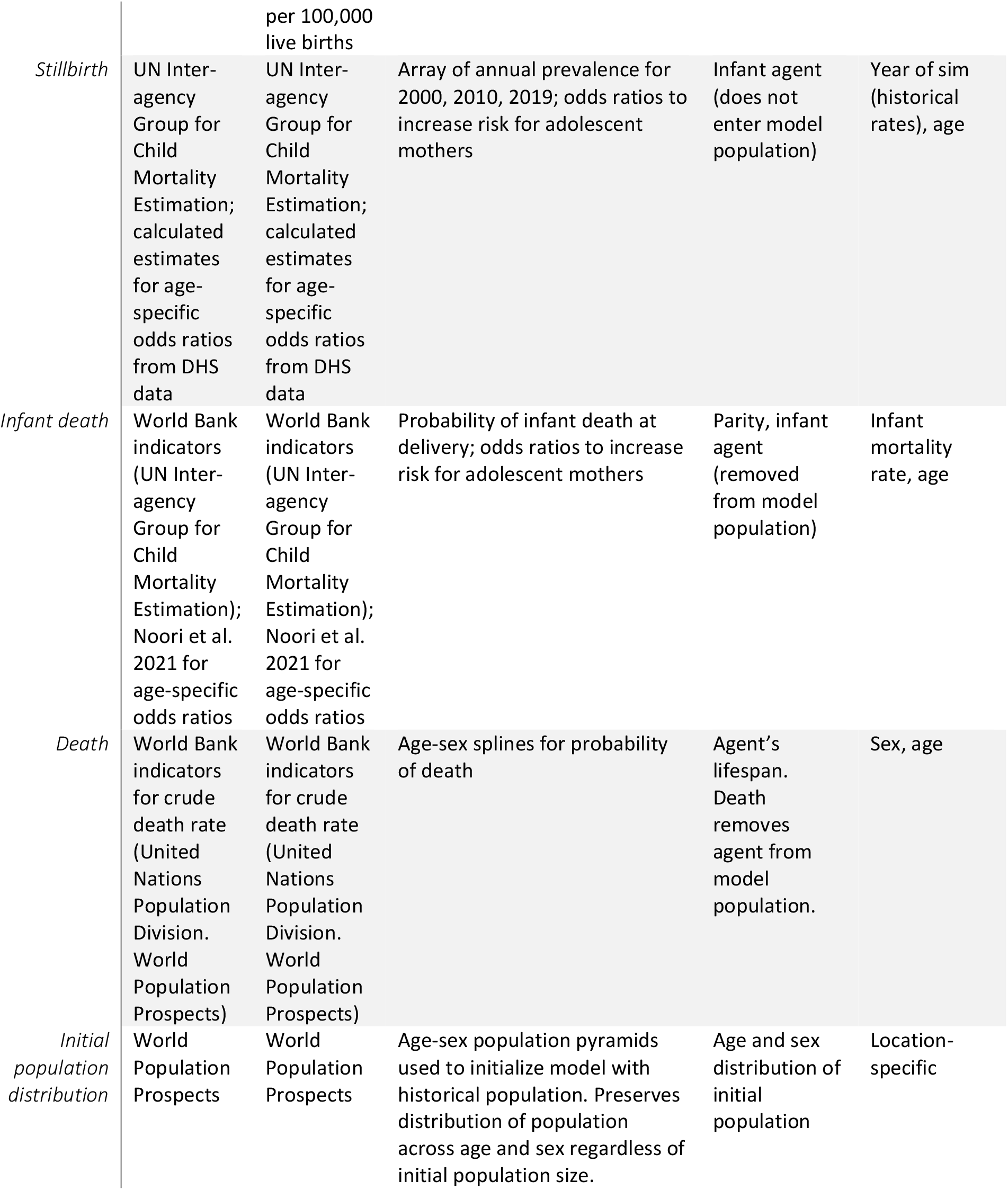
List of parameters, events, and respective data sources in FPsim

## Notes

### Competing Interest Statement

All authors are paid employees of the Bill & Melinda Gates Foundation.

### Funding Statement

This study did not receive any funding.

### Author Declarations

The study used only openly available human data originally derived from the Demographic and Health Surveys (DHS; dhsprogram.com) and Performance Monitoring in Action (PMA; pmadata.org) projects for the purpose of parameterizing the agent-based model. All output presented is simulated.

